# Favipiravir versus Arbidol for COVID-19: A Randomized Clinical Trial

**DOI:** 10.1101/2020.03.17.20037432

**Authors:** Chang Chen, Yi Zhang, Jianying Huang, Ping Yin, Zhenshun Cheng, Jianyuan Wu, Song Chen, Yongxi Zhang, Bo Chen, Mengxin Lu, Yongwen Luo, Lingao Ju, Jingyi Zhang, Xinghuan Wang

## Abstract

**Background:** No clinically proven effective antiviral strategy exists for the epidemic Coronavirus Disease 2019 (COVID-19).

**Methods:** We conducted a prospective, randomized, controlled, open-label multicenter trial involving adult patients with COVID-19. Patients were randomly assigned in a 1:1 ratio to receive conventional therapy plus Umifenovir (Arbidol) (200mg*3/day) or Favipiravir (1600mg*2/first day followed by 600mg*2/day) for 10 days. The primary outcome was clinical recovery rate of Day 7. Latency to relief for pyrexia and cough, the rate of auxiliary oxygen therapy (AOT) or noninvasive mechanical ventilation (NMV) were the secondary outcomes. Safety data were collected for 17 days.

**Results:** 240 enrolled COVID-19 patients underwent randomization; 120 patients were assigned to receive Favipiravir (116 assessed), and 120 to receive Arbidol (120 assessed). Clinical recovery rate of Day 7 does not significantly differ between Favipiravir group (71/116) and Arbidol group (62/120) (P=0.1396, difference of recovery rate: 0.0954; 95% CI: -0.0305 to 0.2213). Favipiravir led to shorter latencies to relief for both pyrexia (difference: 1.70 days, P<0.0001) and cough (difference: 1.75 days, P<0.0001). No difference was observed of AOT or NMV rate (both P>0.05). The most frequently observed Favipiravir-associated adverse event was raised serum uric acid (16/116, OR: 5.52, P=0.0014).

**Conclusions:** Among patients with COVID-19, Favipiravir, compared to Arbidol, did not significantly improve the clinically recovery rate at Day 7. Favipiravir significantly improved the latency to relief for pyrexia and cough. Adverse effects caused Favipiravir are mild and manageable. This trial is registered with Chictr.org.cn (ChiCTR2000030254).

## Introduction

The epidemic coronavirus disease 2019 (COVID-19) caused by SARS-CoV-2 started in Dec. 2019. As of Apr. 8, 2020, the WHO reported 1,452,178 confirmed cases across more than 130 countries with a global mortality rate of 5.8%.^1,2^

At the moment, no specific treatment exists for COVID-19. Standard practice of care focus on treating the clinical symptoms (pyrexia, cough, and acute respiratory distress syndromes (ARDS)) of patients with supportive care such as fluid management and auxiliary oxygen therapy. No proven clinical efficacy of antiviral agent for COVID-19 were reported, whilst some of them (Remdesivir, hINFa-2b, Ribavirin, Chloroquine and Arbidol) are currently under clinical trials for COVID-19.^3,4^

SARS-CoV-2 and influenza viruses exhibit similar disease presentations with similar organ trophism. Because both viruses are RNA virus depending on RNA-dependent RNA polymerase (RdRp) to replicate, the RdRp inhibitor Arbidol (common name for Umifenovir) approved for influenza in Russia and China has been proposed as a standard care option for COVID-19, mainly based on its mechanism-of-action (MoA) and its effects in treating influenza-associated pneumonia.^5-7^

Favipiravir, an antiviral drug targeting RdRP,^8^ approved in Japan for influenza, has an IC50 of 0.013-0.48 ug/ml for influenza A. Comparing this with the EC50 of 2.7-13.8 ug/ml of Arbidol,^9^ we consider Favipiravir might serve as a potential candidate to treat COVID-19. Specifically, we hypothesized that Favipiravir would be superior to Arbidol in terms of improving clinical recovery rate at Day 7, and alleviating pyrexia, cough, and ARDS. To evaluate the clinical efficacy and safety of Favipiravir versus Arbidol as treatment for COVID-19, we conducted a prospective, randomized, controlled, open-label multicenter trial, in adult patients with COVID-19.

## Methods

### Patients

Patients were assessed for eligibility on the basis of: (1) aged 18 years or older; (2) voluntarily provided informed consent; (3) initial symptoms were within 12 days; (4) Diagnosed as COVID-19 pneumonia. According to the Chinese Diagnosis and Treatment Protocol for Novel Coronavirus Pneumonia at that moment,^10,11^ COVID-19 could be diagnosed without a positive SARS-CoV-2 nucleic acid test result by: (1) a positive chest CT scan; (2) significant clinical manifestation including pyrexia, cough, breath difficulty and other indications of viral infection of lower respiratory tract; and (3) laboratory results indicating lymphopenia and (optional) leukopenia. Hence, male and female adult patients with clinically confirmed COVID-19 including moderate, severe or critical types of COVID-19 were eligible. Patients were excluded if they meet any of following criteria: (1) allergic to Favipiravir or Arbidol; (2) with elevated ALT/AST (>6x upper limit of normal range) or with chronic liver disease (cirrhosis at grade Child-Pugh C); (3) severe/critical patients whose expected survival time were <48 hours; (4) female in pregnancy; (5) HIV infection; or (6) considered unsuitable by researchers for the patient’s best interest. Written informed consents were obtained from all patients or their authorized representatives if the patient was unable to write physically.

### Study design

The study was designed as a prospective, randomized, controlled, open-label multicenter trial (Figure 1) conducted from Feb. 20 to Mar. 1, 2020 in three hospitals (Zhonghan Hospital of Wuhan University (ZNWU), Leishenshan Hospital (LSS) and the Third Hospital of Hubei Province (HBTH)) of Wuhan, Hubei, China. A total of 240 patients with COVID-19 pneumonia were recruited from the three hospitals (120 from ZNWU, 88 from LSS, and 32 from HBTH). Patients receive either Favipiravir (1600mg, twice first day followed by 600mg, twice daily, for the following days) or Arbidol (200mg, three times daily) plus standard care for 7 days. The treatment could be extended to 10 days at judgement of the researchers. Standard care could comprise, as for the patient’s best interest, traditional Chinese herbal medicine, antibiotics, additional antiviral treatment, immunomodulatory drugs, steroids, psychotic drugs, nutrition support, cardiovascular drugs, supportive oxygen, noninvasive positive pressure ventilation (NPPV) or invasive ventilation. Randomized open label (1:1 ratio between Arbidol and Favipiravir) was produced by professional statistical software SAS (version 9.4) and assigned to patients. The study was approved by the Institutional Ethics Committee (No. 2020040) (additional details in Protocol/SAP).

**Figure 1.**
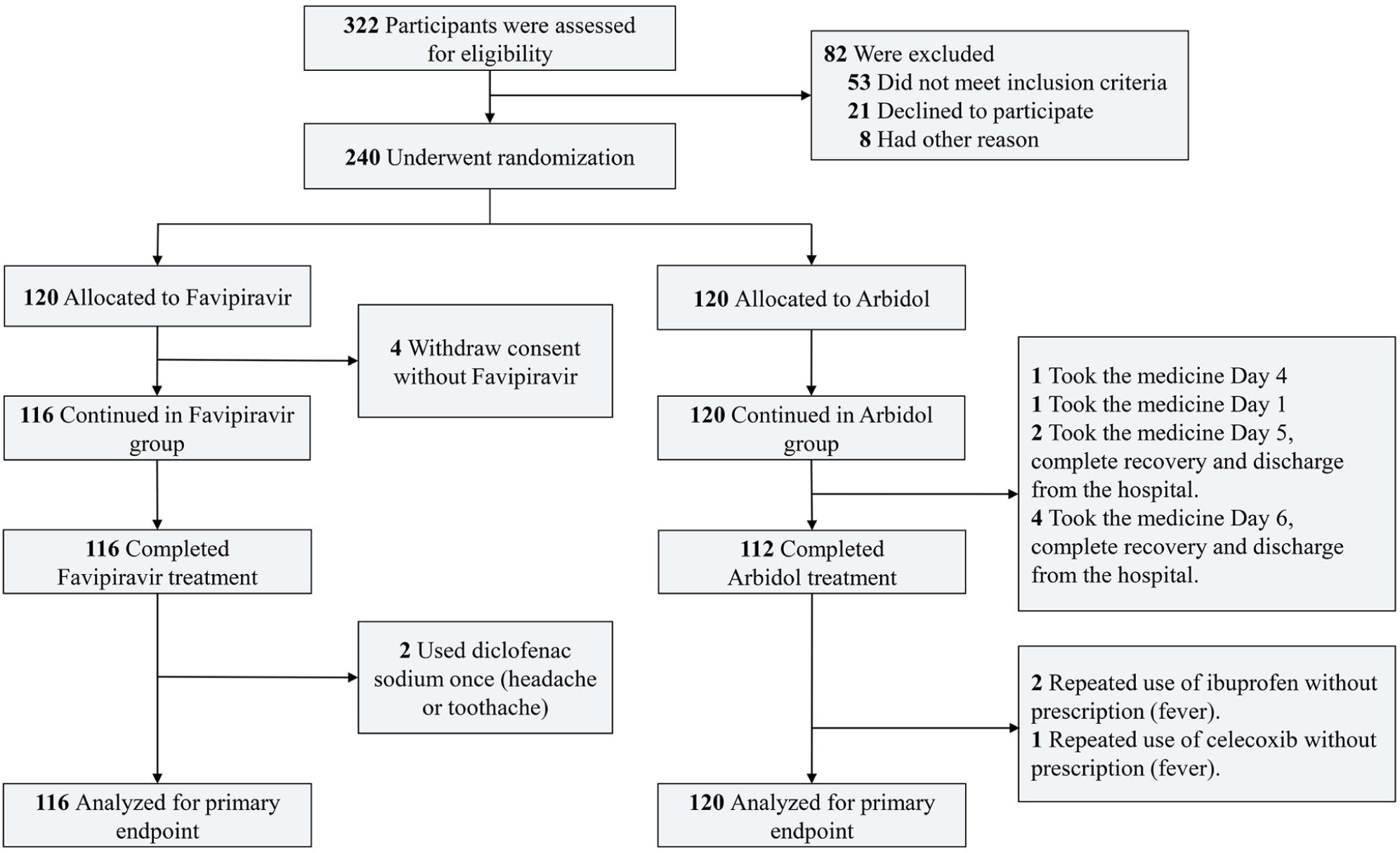
Flow diagram of the study.

### Measurements

Patients were assessed at the time of enrollment for basic physical parameters, body temperature, chronic viral co-infections, HCG (for childbearing age female), COVID-19 clinical classification, SPO2, chest CT, IL-6, blood biochemistry, urinalysis, coagulation function, C-reactive protein and SARS-CoV-2 nucleic acid (additional details in Protocol/SAP). Clinical classification of moderate, severe and critical COVID-19 patients were performed according to the Chinese guideline.^11^ After enrollment, blood biochemistry, urinalysis, coagulation function, C-reactive protein and SARS-CoV-2 nucleic acid were examined on the third (D3±1 day) and the seventh day (D7±2 day) with additional chest CT on the seventh day. The axillary temperature, respiratory rate, oxygen saturation without supportive oxygen, usage of oxygen therapy and noninvasive positive pressure ventilation (NPPV) were recorded in daily follow-up. Repeated measurements were made at least twice in each follow-up. The measurements were taken after 15 minutes rest at room temperature (23±2°C). Adverse events and concomitant medication were observed.

### Outcome definitions

The primary outcome was the clinical recovery rate at 7 days from the beginning of treatment. Clinical recovery was defined as continuous (>72 hours) recovery of body temperature, respiratory rate, oxygen saturation and cough relief after treatment, with following quantitative criteria: axillary temperature ≤36.6°C; respiratory frequency ≤24 times/min; Oxygen saturation ≥98% without oxygen inhalation; mild or no cough. Secondary outcomes included the latency to pyrexia relief (for patients with pyrexia at the time of enrollment), the latency to cough relief (for patients with moderate or severe cough at the time of enrollment), the rate of AOT or NMV, all-cause mortality, dyspnea, rate of respiratory failure (defined as SPO_2_ ≤90% without oxygen inhalation or PaO_2_/FiO_2_ <300mmHg, requires oxygen therapy or additional respiratory support), and the rate of patients needed to receive intensive care in ICU. Safety outcomes included adverse events occurred during treatment and premature discontinuation.

### Statistical analysis

With a limited knowledge of the efficacy for Arbidol, we assumed a 50% clinical recovery rate at Day 7 for the Arbidol group. A superior clinically efficacy of Favipiravir was then expected to be at least 70%. With α=0.025 (single side), β=0.20, power=0.80 we estimated 92 participants was required for each group. The sample size increased about 20% considering shedding/elimination. Hence, the trial was designed to include 240 participants in the group, including 120 in the experimental group and 120 in the control group. SAS (v9.4) software was used for statistical analysis. For primary outcome (clinical recovery rate at Day 7), the comparison between the experimental group and the control group adopts the optimal test. Bilateral 95% CI of the difference between the clinical recovery rate of the experimental group and the control group was calculated. If the lower limit was larger than 0, it was considered the experimental group is superior to the control group. Log rank test was used to compare the recovery latency between the two groups. For secondary outcomes, student’s T-test or Wilcoxon rank sum test (if T-test was not applicable) was performed for safety indicators and continuous variables, Wilcoxon rank sum test was used for grade variables. Frequency or composition (%) were used for statistical description of categorial variables, and Chi-square test or Fisher’s exact test was used for comparison between groups. For all statistical tests, P<0.05 (bilateral) were considered as statistically significant.

## Results

### Clinical characteristics of patients

A total of 240 patients with COVID-19 were enrolled, in which 236 took at least one dosage of drug were considered as the full analysis set (FAS) (Figure S1-S2). The FAS set includes 116 in the Favipiravir group and 120 in the Arbidol group (Table 1). In the Favipiravir group, 59 (50.86%) were males and 57 (49.14%) were females, 87 (75.00%) were <65 years and 29 (25.00%) were ≥65 years, 36 (31.03%) were with hypertension and 14 (12.07%) with diabetes. In the Arbidol group, 51 (42.50%) were males and 69 (57.50%) were females, 79 (65.83%) were <65 years and 41 (34.17%) were ≥65 years, 30 (25.00%) were with hypertension and 13 (10.83%) with diabetes. Main signs and symptoms for enrolled patients were pyrexia, fatigue, dry cough, myalgia, dyspnea, expectoration, sore throat, diarrhea, dizziness, insomnia and conjunctivitis, none of which were significantly different between groups. There was no difference between the time from onset of patient symptoms to time of treatment initiation between groups. Neither the SARS-CoV-2 nucleic acid tests, lymphocyte count, erythrocyte sedimentation rate and C-reactive protein differed between groups (Table 1). 116 cases in Favipiravir group and 119 in Arbidol group underwent chest CT, in which 112 (96.55%) and 114 (95.80%) were diagnosed COVID-19 pneumonia according to the diagnostic criteria (P=0.7635). Overall, no significant difference of basic characteristics of patients between the two groups was observed. However, we noticed a marginally increased ratio of severe to critical patients in the Favipiravir group (16 (severe)+2 (critical)) compared to Arbidol group (8+1) (P=0.0658, Fisher’s exact test, OR: 2.25 [0.91-5.98]).

**Table 1.**
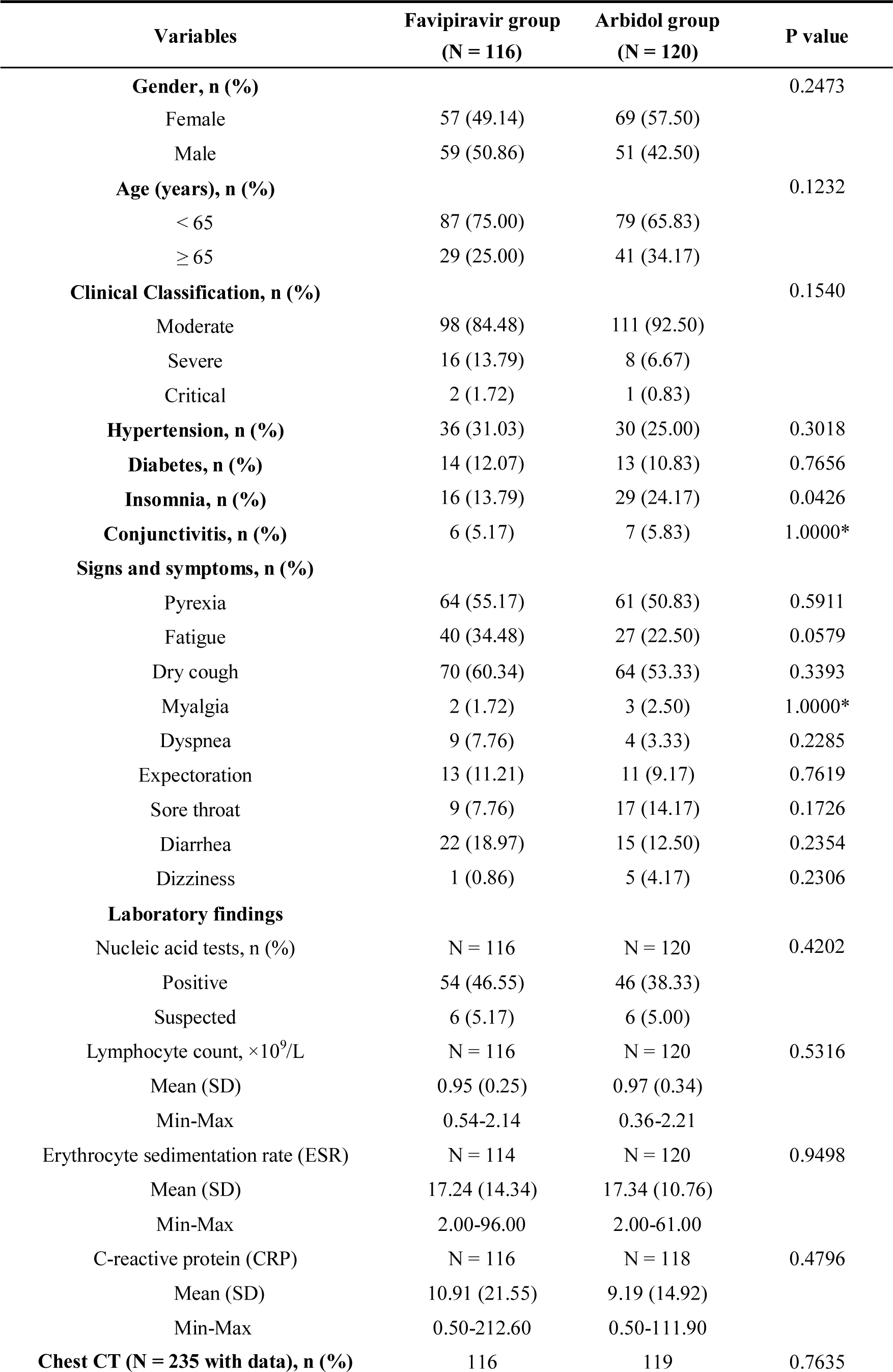
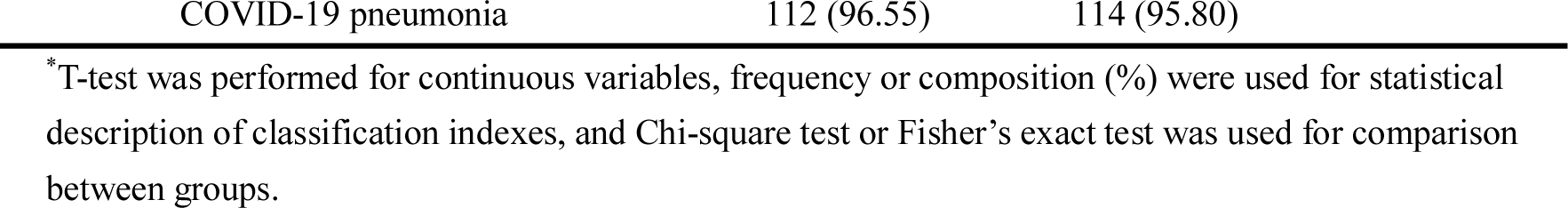
Basic characteristics of the participants.

| **Variables** | **Favipiravir group**<br>**(N = 116)** | **Arbidol group**<br>**(N = 120)** | **P value** |
| --- | --- | --- | --- |
| **Gender, n (%)** |  |  | 0.2473 |
| Female | 57 (49.14) | 69 (57.50) |  |
| Male | 59 (50.86) | 51 (42.50) |  |
| **Age (years), n (%)** |  |  | 0.1232 |
| < 65 | 87 (75.00) | 79 (65.83) |  |
| ≥ 65 | 29 (25.00) | 41 (34.17) |  |
| **Clinical Classification, n (%)** |  |  | 0.1540 |
| Moderate | 98 (84.48) | 111 (92.50) |  |
| Severe | 16 (13.79) | 8 (6.67) |  |
| Critical | 2 (1.72) | 1 (0.83) |  |
| **Hypertension, n (%)** | 36 (31.03) | 30 (25.00) | 0.3018 |
| **Diabetes, n (%)** | 14 (12.07) | 13 (10.83) | 0.7656 |
| **Insomnia, n (%)** | 16 (13.79) | 29 (24.17) | 0.0426 |
| **Conjunctivitis, n (%)** | 6 (5.17) | 7 (5.83) | 1.0000* |
| **Signs and symptoms, n (%)** |  |  |  |
| Pyrexia | 64 (55.17) | 61 (50.83) | 0.5911 |
| Fatigue | 40 (34.48) | 27 (22.50) | 0.0579 |
| Dry cough | 70 (60.34) | 64 (53.33) | 0.3393 |
| Myalgia | 2 (1.72) | 3 (2.50) | 1.0000* |
| Dyspnea | 9 (7.76) | 4 (3.33) | 0.2285 |
| Expectoration | 13 (11.21) | 11 (9.17) | 0.7619 |
| Sore throat | 9 (7.76) | 17 (14.17) | 0.1726 |
| Diarrhea | 22 (18.97) | 15 (12.50) | 0.2354 |
| Dizziness | 1 (0.86) | 5 (4.17) | 0.2306 |
| **Laboratory findings** |  |  |  |
| Nucleic acid tests, n (%) | N = 116 | N = 120 | 0.4202 |
| Positive | 54 (46.55) | 46 (38.33) |  |
| Suspected | 6 (5.17) | 6 (5.00) |  |
| Lymphocyte count, ×10<sup>9</sup>/L | N = 116 | N = 120 | 0.5316 |
| Mean (SD) | 0.95 (0.25) | 0.97 (0.34) |  |
| Min-Max | 0.54-2.14 | 0.36-2.21 |  |
| Erythrocyte sedimentation rate (ESR) | N = 114 | N = 120 | 0.9498 |
| Mean (SD) | 17.24 (14.34) | 17.34 (10.76) |  |
| Min-Max | 2.00-96.00 | 2.00-61.00 |  |
| C-reactive protein (CRP) | N = 116 | N = 118 | 0.4796 |
| Mean (SD) | 10.91 (21.55) | 9.19 (14.92) |  |
| Min-Max | 0.50-212.60 | 0.50-111.90 |  |
| **Chest CT (N = 235 with data), n (%)** | 116 | 119 | 0.7635 |

|  |  |  |
| --- | --- | --- |
| COVID-19 pneumonia | 112 (96.55) | 114 (95.80) |

\*T-test was performed for continuous variables, frequency or composition (%) were used for statistical description of classification indexes, and Chi-square test or Fisher's exact test was used for comparison between groups.

### Comparison of clinical recovery rate of Day 7 of Favipiravir and Arbidol in COVID-19 patients

The group statistics of primary and secondary outcomes were presented in Table 2 and Table S1. At Day 7, 62/120 (51.67%) in the Arbidol group and 71/116 (61.21%) patients in the Favipiravir group (P=0.1396) were clinically recovered (difference of recovery rate (DRR): 0.0954, 95% confidence interval (CI): -0.0305∼0.2213) (Figure 2). Hence, we conclude that Favipiravir does not show superior efficacy compared to Arbidol in terms of improving the clinical recovery rate at Day 7.

**Table 2.** Comparison of clinical recovery rate of Day 7.

| **Variables** | **Favipiravir group** | **Arbidol group** | **Rate ratio (95% CI)** | **P value** |
| --- | --- | --- | --- | --- |
| **Total patients** | **(N = 116)** | **(N = 120)** |  | 0.1396 |
| Recovered, n (%) | 71 (61.21) | 62 (51.67) | 0.0954 (-0.0305, 0.2213) |  |
| **Moderate patients** | **(N = 98)** | **(N = 111)** |  |  |
| Recovered, n (%) | 70 (71.43) | 62 (55.86) | 0.1557 (0.0271, 0.2843) | 0.0199 |
| **Severe or critical patients** | **(N = 18)** | **(N = 9)** |  |  |
| Recovered, n (%) | 1 (5.56) | 0 (0.00) | 0.0556 (-0.0503, 0.1614) | 0.4712 |
| **Patients with hypertension and/or diabetes** | **(N = 42)** | **(N = 35)** |  |  |
| Recovered, n (%) | 23 (54.76) | 18 (51.43) | 0.0333 (-0.1904, 0.2571) | 0.7704 |

**Figure 2.**
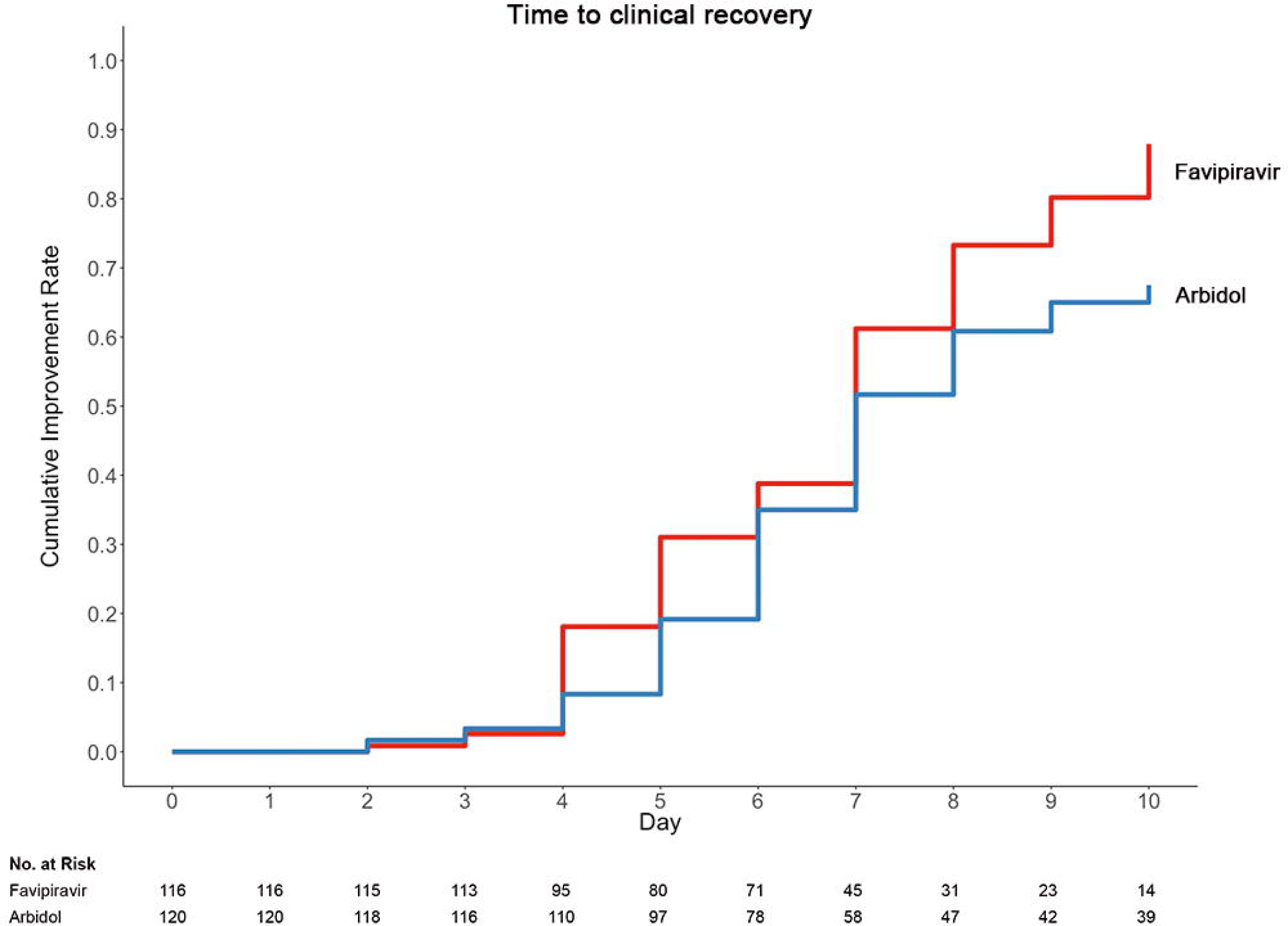
Time to clinical recovery in the trial population.

Post-hoc test for interaction between treatment and clinical classification showed no interaction between these two factors, both of which contributed to the primary outcome (P=0.017 for treatment, and P<0.001 for clinical classification, with a general linear model). A post-hoc analysis found that for moderate patients with COVID-19, clinical recovery of Day 7 was 62/111 (55.86%) in the Arbidol group and 70/98 (71.43%) in the Favipiravir group (P=0.0199) (DRR: 0.1557, 95% CI: 0.0271∼0.2843); for severe/critical patients, clinical recovery rate was 0/9 (0%) in the Arbidol group and 1/18 (5.56%) in the Favipiravir group (P=0.4712) (DRR: 0.0556, 95% CI: -0.0503∼0.1614).

### Comparison of duration of pyrexia, cough relief time and auxiliary oxygen therapy or noninvasive mechanical ventilation rate

Table 3 displayed duration of pyrexia, cough relief time and auxiliary oxygen therapy or noninvasive mechanical ventilation rate between the Favipiravir and Arbidol groups. At baseline, for patients in Favipiravir group, 71/116 (61.2%) had pyrexia and 78/116 (67.2%) had cough; for patients in the Arbidol group, 74/120 (61.7%) had pyrexia and 73/120 (60.8%) had cough. Whilst the incidence of pyrexia and cough did not differ between two groups at baseline, both the latency to pyrexia reduction and cough relief in the Favipiravir group was significantly shorter than that in the Arbidol group (P<0.0001) (Figure 3).

**Table 3.** Comparison of time to relief for pyrexia, cough relief time and other secondary outcomes.

| Variables | Time to relief for pyrexia |  | Cough relief time |  |
| --- | --- | --- | --- | --- |
|  | Favipiravir group | Arbidol group | Favipiravir group | Arbidol group |
| Total patients | (N = 71) | (N = 74) | (N = 78) | (N = 73) |
| Day 1 | 15 (21.13) | 2 (2.70) | 1 (1.28) | 3 (4.11) |
| Day 2 | 23 (32.39) | 8 (10.81) | 2 (2.56) | 1 (1.37) |
| Day 3 | 19 (26.76) | 18 (24.32) | 23 (29.49) | 7 (9.59) |
| Day 4 | 10 (14.08) | 15 (20.27) | 20 (25.64) | 11 (15.07) |
| Day 5 | 1 (1.41) | 16 (21.62) | 10 (12.82) | 12 (16.44) |
| Day 6 | - | 5 (6.76) | 10 (12.82) | 10 (13.70) |
| Day 7 | - | 3 (4.05) | 3 (3.85) | 3 (4.11) |
| Day 8 | - | - | 7 (8.97) | 6 (8.22) |
| Day 9 | - | - | 1 (1.28) | 3 (4.11) |
| Censored | - | - | 1 (1.28) | 17 (23.29) |
| Log-rank P value | < 0.0001 |  | < 0.0001 |  |

| Other secondary outcomes |  |  |  |  |
| --- | --- | --- | --- | --- |
| AOT or NMV* | Favipiravir group | Arbidol group | Rate ratio (95% CI) | P value |
| Total patients | N = 116 | N = 120 |  |  |
| With auxiliary, n (%) | 21 (18.10) | 27 (22.50) | -0.0440 (-0.1464, -0.0585) | 0.4015 |
| Patients with hypertension and/or diabetes | N = 42 | N = 35 |  |  |
| With auxiliary, n (%) | 9 (21.43) | 10 (28.57) | -0.0714 (-0.2658, 0.1230) | 0.4691 |
| All-cause mortality | 0 (0.00) | 0 (0.00) | / | / |
| Dyspnea after taking medicine, n (%) | 4 (3.45) | 14 (11.67) | / | 0.0174 |
| Respiratory failure, n (%) | 1 (0.86) | 4 (3.33) | / | 0.3700* |

\*Fisher's exact test was used for comparison between groups.
AOT: Auxiliary oxygen therapy.
NMV: Noninvasive mechanical ventilation.

**Figure 3.**
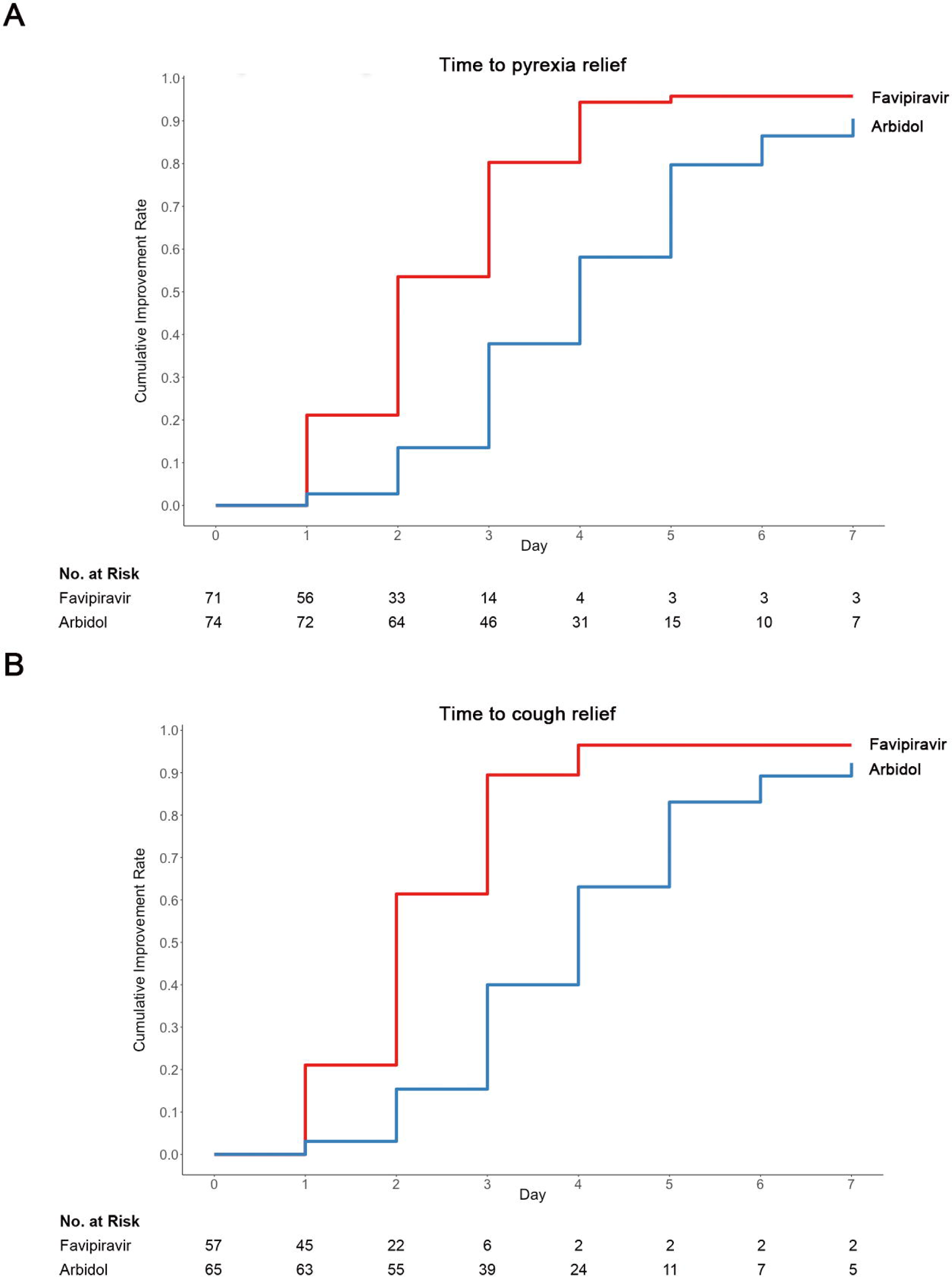
Time to (A) pyrexia or (B) cough relief in the trial population.

The incidence of *de novo* auxiliary oxygen therapy (AOT) or noninvasive mechanical ventilation (NMV) was 27/120 (22.50%) in the Arbidol group and 21/116 (18.10%) in the Favipiravir group (P=0.4015) (DRR: -4.40%, 95% CI: -14.64%∼5.85%). For all cases enrolled in this study, the all-cause mortality was 0. The number of cases of respiratory failure were 4 in Arbidol group and 1 in Favipiravir group (P=0.3700). Patients with dyspnea was 15/120 (12.5%) in the Arbidol group and 13/116 (11.2%) in the Favipiravir group (P=0.7588). A post-hoc analysis showed that *de novo* incidences of dyspnea during the course of treatment occurred at 4/116 (3.45%) patients in the Favipiravir group and 14/120 (11.67%) patients in the Arbidol group (P=0.0174). Hence, we conclude that for secondary outcomes, Favipiravir significantly shortened the latency to relief for cough and pyrexia. Favipiravir was not associated with a decreased rate for AOT or NMV, dyspnea, overall respiratory failure rate, ICU admission or all-cause mortality.

### Comparison of antiviral-associated adverse effects

During this trial, we detected 37 incidences of antiviral-associated adverse effects (AE) in the Favipiravir group and 28 incidences in the Arbidol group. All observed AE incidences were level 1. Favipiravir was associated with increased serum uric acid (3 (2.50%) in Arbidol group vs 16 (13.79%) in Favipiravir group, P=0.0014). No statistical difference was observed for the frequency of abnormal ALT/AST, psychiatric symptom reactions or digestive tract reactions (Table 4). Most of these adverse reactions disappeared by the time patients being discharged.

**Table 4.**
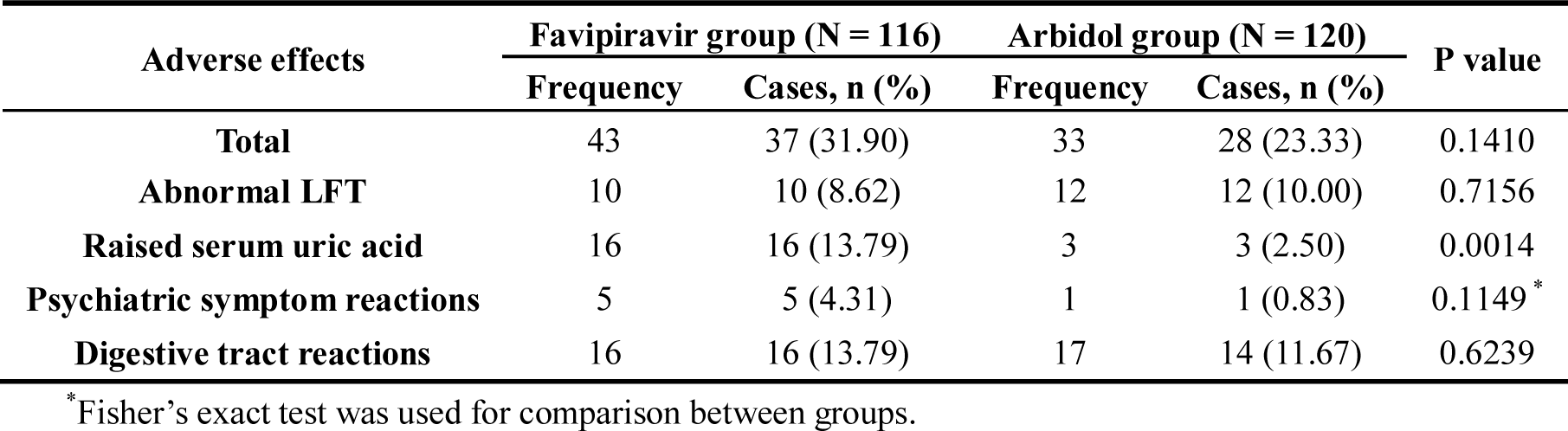
Comparison of antiviral-associated adverse effects.

| Adverse effects | Favipiravir group (N = 116) |  | Arbidol group (N = 120) |  | P value |
| --- | --- | --- | --- | --- | --- |
|  | Frequency | Cases, n (%) | Frequency | Cases, n (%) |  |
| **Total** | 43 | 37 (31.90) | 33 | 28 (23.33) | 0.1410 |
| **Abnormal LFT** | 10 | 10 (8.62) | 12 | 12 (10.00) | 0.7156 |
| **Raised serum uric acid** | 16 | 16 (13.79) | 3 | 3 (2.50) | 0.0014 |
| **Psychiatric symptom reactions** | 5 | 5 (4.31) | 1 | 1 (0.83) | 0.1149 * |
| **Digestive tract reactions** | 16 | 16 (13.79) | 17 | 14 (11.67) | 0.6239 |

\*Fisher's exact test was used for comparison between groups.

## Discussion

We conducted a prospective, multicenter, open-labeled, randomized superiority clinical trial and hypothesized that Favipiravir would be superior to Arbidol in terms of efficacy for moderate symptoms, and would accelerate the clinical recovery of pyrexia, cough, and breathing difficulties compared with Arbidol.

Favipiravir treatment did not improve clinical recovery rate of Day 7 (61.21%) compared to Arbidol group (51.67%). However, it did significantly improve the latency to cough relief and decreased the duration of pyrexia. Favipiravir was not associated with any differences in ICU admission, AOT/NMV, dyspnea, respiratory failure or all-cause mortality.

Interestingly, post-hoc observation showed that a trend of Favipiravir being effective to improve clinical recovery rate at Day 7 in moderate COVID-19 patients compared to Arbidol. This effect diminished for severe/critical COVID-19 patients. Additionally, post-hoc analysis showed that for moderate COVID-19 patients, Favipiravir was associated with decreased auxiliary oxygen therapy or noninvasive mechanical ventilation rate with marginal significance (P=0.0541). Finally, in the FAS, post-hoc analysis also showed that Favipiravir treatment significantly decreased *de novo* incidences of dyspnea. Whether Favipiravir would be only effective for moderate COVID-19 patients, or could Favipiravir be used to prevent disease progression, is a question warrant future investigation.

The combination of traditional Chinese medicine and antiviral drugs is more common in China, which is due to the traditional medical culture background of the treatment of choice. Also, anti-infection and immune regulation play an important role in the treatment of the COVID-19. Ancillary treatments, such as traditional Chinese medicine, anti-infection and immunomodulatory drugs, were without statistical difference between groups (Table S2).

Our trial has several limitations. Firstly, for COVID-19, there is no clinically proven effective antiviral drug to serve as the control arm. Although Chinese guideline had recommended several options including Arbidol,^11^ no RCT results on these drugs were reported. Arbidol was widely used by Chinese doctors in the beginning stage of this epidemic of COVID-19 (Jan. 1-30, 2020) based on *in vitro* evidence.^12^ For ethical reasons, we chose Arbidol for the control arm. Secondly, observation time frame was limited due to the urgency of this epidemic. For the same reason, no relapse (including nucleic acid conversion, pyrexia, cough, or pneumonia progress by radiology) tracking were performed for the discharged patients. Thirdly, in the inclusion criteria, we did not force positive nucleic acid test as a necessity. The accuracy of nucleic acid assay was limited, which might due to multiple reasons including previous treatment, latency of onset, sampling method, biological specimen characteristics. This particular accuracy problem was a known issue among clinical practitioners across the world. It was estimated that the assay might have at most 30%-50% of sensitivity for patients in early stage of the disease, whilst contact history, clinical manifestations, radiology evidences, and lab results including leukopenia and lymphopenia could be confirmatory for these nucleic-acid-negative pneumonia patients. In the Chinese guideline,^11^ patients meeting these criteria were considered as with clinically confirmed COVID-19. In this trial, 46.55% patients in Favipiravir group and 38.33% in Arbidol group were nucleic-acid-positive at enrollment. Considering the population incidence of COVID-19 infection at the time of this trial in Wuhan, we consider the probability for mis-identifying patients of pneumonia disease other than COVID-19 into this trial is low. Fourthly, the protocol does not prespecify clinical classification as a stratification factor. Ethical concerns arose against completely excluding severe/critical cases from potential beneficial treatment. Additionally, because of the complexity of the disease, progression from moderate to severe/critical is possible. Terminating trial treatment to such patients from the study was considered unacceptable. Post-hoc analysis showed that both treatment and clinical classification contributed significantly to the primary outcome of clinical recovery rate at Day 7. Difference of the frequency of severe/critical patients between groups reached a marginal significance, which made an important impact on the trial outcome.

## Conclusions

Compared to Arbidol, Favipiravir does not significantly improve clinical recovery rate of Day 7. Favipiravir is associated with significantly shortened latency to relief for pyrexia and cough. Antiviral-associated adverse effects of Favipiravir are mild and manageable.

## Data Availability

With the permission of the corresponding author, we can provide participant data, statistical analysis.

## Funding and disclosures

This work was supported by the National Key Research and Development Program of China (2020YFC0844400). The funder had no role in the design and conduct of the study; collection, management, analysis, and interpretation of the data; preparation, review, or approval of the manuscript; and decision to submit the manuscript for publication. All authors have no conflicts of interest to declare.

We would like to acknowledge the support of China National Center for Biotechnology Development, Science and Technology Department of Hubei Province. We thank the excellent technical assistance of Dr. Xiuli Zhao (Beijing Tongren Hospital, Capital Medical University, Beijing, China). We also thank the help from the rescue medical teams for Hubei province (Tianjin medical team, Liaoning medical team, Guangdong Province Traditional Chinese Medical Hospital, Longhua Hospital Shanghai University of Traditional Chinese Medicine, Liyuan Hospital of Tongji Medical College of Huazhong University of Science and Technology, and Puren Hospital of Wuhan University of Science and Technology).

## Notes

### Competing Interest Statement

The authors have declared no competing interest.

### Clinical Trial

This trial is registered with Chictr.org.cn (ChiCTR2000030254).

